# Thermal Discomfort and *In-vitro* Fertilization Treatment Outcomes

**DOI:** 10.64898/2026.05.21.26353705

**Authors:** Venkata Madhavi Latha Telagarapu, Sriharsha Ravuri, Padmaja Veeramachaneni, Sireesha R. Bankuru, Naresh Kumar

## Abstract

**Background:** Literature on the role of thermal discomfort (heat- and cold-stress) on in-vitro fertilization (IVF) outcomes are scarce and inconclusive. This multi-center research examines association between heat stress and IVF treatment outcomes in Andhra Pradesh, which is prone to year around chronic heat stress.

**Methods:** IVF data were abstracted from clinical chart review of all patients from three IVF centers from 2019 to 2023, which included time-stamped data on each IVF procedure, demographics and pre-existing comorbidities. Weather data were acquired from the National Climatic Data Center (NCDC). IVF outcomes were modelled with respect to time-lagged exposure to ambient temperature stratified by hyper- and hypothermic conditions using Poisson and logistic regressions depending on the scale of IVF outcomes adjusting for confounders.

**Results:** A total of 690 unique patients had 1,311 encounters at the three IVF clinics between 2019 and 2023. Oocytes were retrieved 717 times and fertilized by ICSI. Embryo transfers (ET) were conducted 978 times. On average, ET was attempted 1.42 times per patient. Of 978 ET attempts, clinical pregnancy was confirmed for 249(25.46%) by doing an ultrasound. Of the confirmed clinical pregnancies, 136 (45.37%) ended spontaneous abortions/miscarriage (SAM). Heat stress peaked in June, which corresponded with elevated number of SAM. Under hypo- and hyper-thermic conditions a unit increase ambient temperature was associated with an 11% higher and an 8% lower number of oocytes retrieved, respectively. Adjusting for confounders, a 10° F increase in two-day lag heat stress was associated with a 30% higher odds of SAM (odds ratio ~ 1.03; 95% CI = 1.001 to 1.068; p-value < 0.043), and odds of PTB were 3 times higher when three day-lagged heat index (HI) was greater than 35° C (odds ratio 1.13 to 7.99; p < 0.05).

**Conclusion:** Our findings warrant strategies to engage IVF patients in mitigating their exposure to thermal discomfort and thermal shock before and during the IVF treatment.

## A. INTRODUCTION

Global fertility has been declining for the past several decades, with the total fertility rate (TFR) declining from 4.8 in 1950 to 2.2 in 2021.^1^ Moreover this downward trend is projected worldwide decreasing below the replacement rate of 2.1.^2^ While demographic shifts, including delayed parenthood, rising maternal age, and socioeconomic factors, such as increasing costs of raising children, are important drivers of the declining fertility rates, higher infertility levels are also contributing to this declining trend.^3,4^ Approximately one in six adults (i.e. ~ 17.5% of the global adult population) are affected by infertility at some point in their lives,^5^ resulting in an increased demand for infertility treatment. *In-vitro* fertilization (IVF) is an advanced assisted reproductive technique and its demand has been rising, e.g. IVF demand increased from 2.5 million cycles in 2019^6^ to more than 3 million in 2022,^7^ assisting millions of couples to have children.^8^

IVF treatment is a multi-stage and complex process and many factors affect the treatment, including patients’ medical conditions, demographics, therapeutic interventions, lifestyle and environmental stressors (ES).^9,10^ ES are of particular interest not only because ES are intensifying due to changing climate and extreme weathers but also because ES exposures are avoidable and modifiable. Emerging literature suggests that exposure to ES, including heat stress and persistent pollutants, affect endocrine functions and damage reproductive health in both males and females.^11,12^ Among ES, this paper will focus on the effects of heat stress on IVF outcomes at different stages ranging from growth and development of oocytes to pregnancy outcomes^13,14^. Heat stress initiates cytoplasmic responses, triggering the transcription of heat shock proteins (HSPs),^15^ which act as chaperones and protect cells from ES induced damage and maintain cellular homeostasis^16^ by inducting pro-inflammatory and anti-inflammatory cytokines, folding of newly synthesized proteins and degradation of unstable or misfolded proteins.^17,18^ Heat stress also activates reactive oxygen species (ROS) causing a variety of cellular changes, including cytoskeletal reorganization, chromatin precipitation, organelle migration, and mitochondrial edema, disrupting chromosome separation and impacting cellular functions.^19^ Heat stress is particularly relevant for IVF, as its stress impairs the reproductive ability of females, including ovarian follicle formation in utero,^14,20^ oocyte disruption and embryo development via apoptosis caused by impaired mitochondrial DNA transcription.^20^ Heat stress also induces cytoplasmic alterations in oocytes and early embryos, including cytoskeletal reorganization, chromatin condensation, abnormal organelle migration, mitochondrial edema, and disruptions in chromosome segregation, collectively reducing oocyte maturation, embryo quality, and implantation potential.^21^ Dysfunction in maternal mitochondria is also inherited by embryos, which impairs essential energy supply during early embryonic divisions,^22^ influencing subsequent stages of the IVF process including embryo implantation, development of the fetus and placenta.^23^ Thus, we will examine the role of time-lagged heat stress and thermal shock (i.e. sudden changes in temperature) in IVF outcomes.

There are limited clinical and/or epidemiological studies on the role of heat stress in IVF, because most of our knowledge on the role of heat stress in reproductive health, including IVF, builds on animal-based toxicological studies.^24^ Clinical/epidemiological studies largely focus on the role of seasonal variations and meteorological conditions in IVF outcomes. Moreover, the findings of these studies have been inconclusive and inconsistent and lack control for potential confounders.^25^ For example, some studies show no significant seasonal differences in the number of oocytes retrieved.^26,27^ However, a study at the IVF-ET unit of Hadassah Hebrew University Medical Centers in Jerusalem found that fertilization rates and embryo quality varied significantly throughout the year with highest in spring and lowest in autumn, correlating with longer daylight hours rather than with temperature or other environmental factors.^28^ Another study explored the role of daily average temperatures in IVF outcomes and reported that exposure to higher temperatures after confirmation of biochemical pregnancy or clinical pregnancy was negatively associated with the live birth rate.^29^ This paper will examine the role of thermal discomfort (both heat- and cold-stress), sudden changes in temperature and seasonality in IVF outcomes, ranging from oocyte quantity to pregnancy outcomes.

## B. METHODS AND MATERIAL

### B1. Study population

This study included women who underwent IVF treatment between 2019 and 2023 at three IVF centers in Andhra Pradesh, South India. We used the following inclusion and exclusion criteria:

#### Inclusion criteria

We included patients who: 1) were 18 years or older and underwent IVF treatment between 2019 and 2023; 2) participated in IVF cycles that followed antagonist protocol for ovarian stimulation; and 3) underwent embryos transfer (ET).

#### Exclusion criteria

Patients were excluded if 1) they cancelled at any phase of the cycle; 2) their data were incomplete; and 3) those used surrogate cycles.

### B2. IVF procedure

All IVF cycles were downregulated before starting the procedure. A transvaginal ultrasonography was performed on the second day of the menstrual cycle to assess antral follicular count and functional cysts at baseline for all patients. Gonadotropins like recombinant FSH and/or highly purified HMG were started for controlled stimulation of the ovaries according to patient’s age, weight, AMH, and AFC on the day of stimulation. Dose was tailored based on ovarian response during the cycle to ensure optimal follicular growth and maturation. Ultrasonography was repeated periodically to monitor the number and growth of the follicles. In all three centers, controlled ovarian stimulation was performed using a flexible GnRH antagonist protocol, as it poses a lower risk of ovarian hyperstimulation syndrome (OHSS) as compared to long agonist protocol.^30^ GnRH antagonist (cetrorelix) was started when the size of the follicle was 12 to 14 mm or serum estrogen levels were > 400 pg/ml. The treatment continued until the day of ovulation trigger to prevent LH surge, ^31^ avoiding premature ovulation. When at least three or more follicles reached ≥ 17 mm, an injection of human chorionic gonadotropin or GnRH agonist or dual trigger was administered depending on the patient’s medical history, ovarian response, and risk of OHSS to induce final maturation of the eggs within the follicles, preparing them for oocyte retrieval. Approximately 36 hours after ovulation trigger, oocytes were retrieved under general anesthesia. Using visual inspection under the inverted microscope, oocyte morphology and quality were documented as described in Table 1 ^32^.

**Table 1:** Oocyte characterization.

| Stage of oocyte | Maturity | Description |
| --- | --- | --- |
| Germinal Vesicle (GV) | Most Immature | Oocyte with an intracytoplasmic nucleus (meiosis has not yet begun). |
| Metaphase I (MI) | Immature | Oocyte without germinal vesicle and polar body (first phase of meiosis). |
| Metaphase II (MII) | Mature | Oocyte is spherical, enclaved with zona pellucida and has a uniform translucent cytoplasm and 1st polar body (second phase of meiosis). |

Oocytes were then fertilized with sperms with the aid of intracytoplasmic sperm injection (ICSI). After 17–20 h, normal zygotes displaying double pronuclei were identified and subsequently transferred into cleavage fluid for culture, allowing further development of the embryos. Embryos were graded either on the 3^rd^ day or 5^th^ day depending on patient’s age, number of embryos, prior IVF outcomes, embryo developmental progression, and laboratory culture conditions. Embryo quality was assessed following the Istanbul consensus guidelines as: grade 1 = best quality; grade 2 = fair; grade 3 = poor quality.^33^ For some patients, fresh embryos were transferred and remaining viable embryos were frozen for future use. For other patients, all of their viable embryos were frozen for future use.

Two weeks after ET, we tested serum beta HCG and a value ≥ 5 mIU/ml was considered as biochemical pregnancy. Patients with positive biochemical pregnancy were further evaluated after two weeks by transvaginal sonography. Clinical pregnancy was confirmed if they showed one or more gestational sacs or a fetal pole with heartbeat, and those with confirmed pregnancy were followed further. A natural loss of pregnancy within 20 weeks of gestation was considered as a SAM, a live birth with less than 37 weeks of gestation was defined as a per-term birth (PTB), and a live birth > 37 weeks of gestation as a normal birth.

### B3. Data

We used two different data sets: a) de-identified IVF data from three different clinics, and b) meteorological data. This study was approved by the University of Miami Institutional Review Board (IRB protocol number: 20170855).

#### B3.1 IVF data

After reviewing each patients’ IVF charts as detailed above, their time-stamped data were extracted if they met our inclusion criteria. These data included patients’ demographics, height, and weight to calculate BMI, IVF associated medical conditions, anti-Müllerian hormone (AMH) levels, dosage of GnRH antagonists and gonadotropins administered. Our outcome variables were: a) oocytes quantity and quality, b) pregnancy confirmation after embryos transfer (ET), c) SAM, and d) pre-term birth (PTB).

#### B3.2 Meteorological data

Hourly meteorological data, including ambient temperature and relative humidity (RH), were acquired from the National Climatic Data Center (NCDC). These data are publicly available. Given that we needed to compute time-lagged exposure to meteorological condition, we acquired meteorological data three months before and during the study period. We identified all meteorological stations within 0.75° distance (~82.5 km at the equator) from each of the three clinics, assuming patients who underwent IVF treatment in the clinic lived within 0.75 distance from the clinic. We used hourly meteorological data to compute daily average and standard deviation (SD) of temperature, RH, wind velocity and heat index (HI).

### B4. Exposure quantification

Our main causal variables were thermal discomfort and thermal shock, i.e. sudden changes in temperature. We classified ambient temperature under three categories: thermal comfort, defined as temperatures between 24°C and 28°C, heat stress or hyperthermic condition as ambient temperatures ≥ 30°C, and cold stress or hypothermic condition as temperature < 24 °C. We define thermal shock as sudden increase or decrease in temperature. A higher value of SD indicates more thermal shock and vice versa. Our underlying assumption is that human body requires time to acclimatize to changing temperature, and exposure to abrupt changes in temperature will trigger thermal discomfort and induce biophysical changes. Given that there is a delay between exposure to heat- and cold-stress and biophysiological responses, we computed time-lagged exposures prior to each IVF outcome. We computed discrete and distributed time-lagged exposures up to thirty days at 1-d, 3-d and 7-d time intervals. IVF data were integrated with meteorological data based on hospital location and timestamp of these data.

### B5. Statistical Analysis

We analyzed data in STATA (Stata Corp LLC, College Station, TX, USA).^34^ First, we conducted a descriptive analysis to summarize patients’ demographics and outcome variables. Second, we conducted exploratory bivariate analysis, examining associations of each meteorological condition with each IVF outcome. We also computed coefficients of each variable for each lag up to 30 days to examine changes in associations of meteorological conditions with the increase in time lag. Finally, we modeled the risk of different IVF outcomes with respect to time-lagged meteorological conditions, SD of temperature and exposure to thermal discomfort using appropriate regression models, depending on the statistical distribution of the outcome variable, e.g. logistic regression for a binary variable and linear or log-linear regression for continuous outcome variable. We adjusted all models for age, BMI, AMH levels, dose of gonadotropins, and gonadotropin-releasing hormones to address potential confounding biases. For all models a p-value of ≤ 0.05 was considered as statistically significant. The risks of IVF outcomes were reported as odds ratio (OR) or regression coefficient with 95% confidence intervals (CI).

## C. RESULTS

### C1. Sample characteristics

Between January 2019 and December 2023, 690 patients meeting our eligibility criteria were included in this study (Table 2). These patients had 2,379 clinical visits for IVF related procedures, including oocyte retrieval, embryo transfer, pregnancy confirmation by beta HCG and ultrasonography, and SAM. The average age of patients was 28.4 years (95% CI = 27.93 – 28.81) and their body mass index (BMI) was 27.2 kg/m^2^ (95% CI = 26.86 – 27.51).

**Table 2.** Sample composition by clinic, Oocyte retrieval and embryo transfer (ET), 2019–2023.

| Clinic place | # patients | # times oocytes retrieved | # times ET |
| --- | --- | --- | --- |
| Guntur | 285 (41.3) | 286 (39.9) | 343 (34.8) |
| Vijayawada | 224 (32.4) | 248 (34.6) | 402 (40.8) |
| Visakhapatnam | 181 (26.2) | 183 (25.5) | 241 (24.4) |
| Total | 690 (100) | 717 (100) | 986 (100) |

Monthly summary of the outcomes and exposure variables is presented in Table 3. Elevated temperature and heat index were observed between March and August, peaking in May and June. Monthly ambient temperatures ranged from 24.1°C in January to 30.4°C in May and June, and heat index ranged from 76.7°F in January to 98.9°F in June.

**Table 3:**
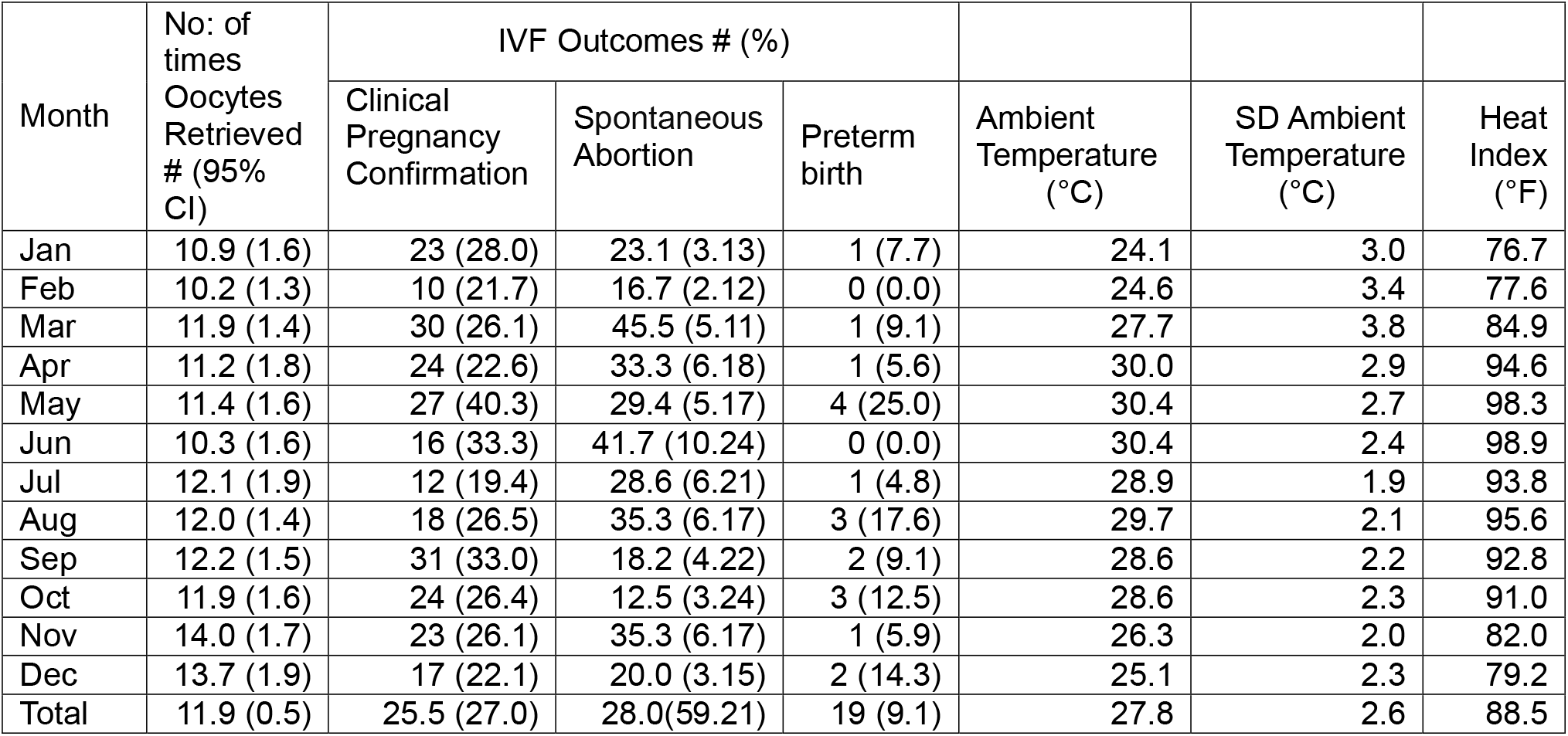
Descriptive statistics of the number of times oocytes retrieved, clinical pregnancy confirmation 4 weeks after embryo transplantation, spontaneous abortion, and meteorological conditions in Andhra Pradesh, India, 2020 – 2023.

| Month | No: of times Oocytes Retrieved # (95% CI) | IVF Outcomes # (%) |  |  | Ambient Temperature (°C) | SD Ambient Temperature (°C) | Heat Index (°F) |
| --- | --- | --- | --- | --- | --- | --- | --- |
|  |  | Clinical Pregnancy Confirmation | Spontaneous Abortion | Preterm birth |  |  |  |
| Jan | 10.9 (1.6) | 23 (28.0) | 23.1 (3.13) | 1 (7.7) | 24.1 | 3.0 | 76.7 |
| Feb | 10.2 (1.3) | 10 (21.7) | 16.7 (2.12) | 0 (0.0) | 24.6 | 3.4 | 77.6 |
| Mar | 11.9 (1.4) | 30 (26.1) | 45.5 (5.11) | 1 (9.1) | 27.7 | 3.8 | 84.9 |
| Apr | 11.2 (1.8) | 24 (22.6) | 33.3 (6.18) | 1 (5.6) | 30.0 | 2.9 | 94.6 |
| May | 11.4 (1.6) | 27 (40.3) | 29.4 (5.17) | 4 (25.0) | 30.4 | 2.7 | 98.3 |
| Jun | 10.3 (1.6) | 16 (33.3) | 41.7 (10.24) | 0 (0.0) | 30.4 | 2.4 | 98.9 |
| Jul | 12.1 (1.9) | 12 (19.4) | 28.6 (6.21) | 1 (4.8) | 28.9 | 1.9 | 93.8 |
| Aug | 12.0 (1.4) | 18 (26.5) | 35.3 (6.17) | 3 (17.6) | 29.7 | 2.1 | 95.6 |
| Sep | 12.2 (1.5) | 31 (33.0) | 18.2 (4.22) | 2 (9.1) | 28.6 | 2.2 | 92.8 |
| Oct | 11.9 (1.6) | 24 (26.4) | 12.5 (3.24) | 3 (12.5) | 28.6 | 2.3 | 91.0 |
| Nov | 14.0 (1.7) | 23 (26.1) | 35.3 (6.17) | 1 (5.9) | 26.3 | 2.0 | 82.0 |
| Dec | 13.7 (1.9) | 17 (22.1) | 20.0 (3.15) | 2 (14.3) | 25.1 | 2.3 | 79.2 |
| Total | 11.9 (0.5) | 25.5 (27.0) | 28.0(59.21) | 19 (9.1) | 27.8 | 2.6 | 88.5 |

### C2. Effect of heat/temperature on oocytes

During the study period, 717 oocyte retrievals were performed. Most of the subjects (95.68%) had oocyte retrieval only once, 3.90% had twice. The average number of oocyte retrievals performed each month ranged from 10.2 to 14.0, with the highest frequency observed in November and the lowest in February (Figure 1). The overall average number of oocytes retrieved per month was 11.9 at each attempt.

**Figure 1:**
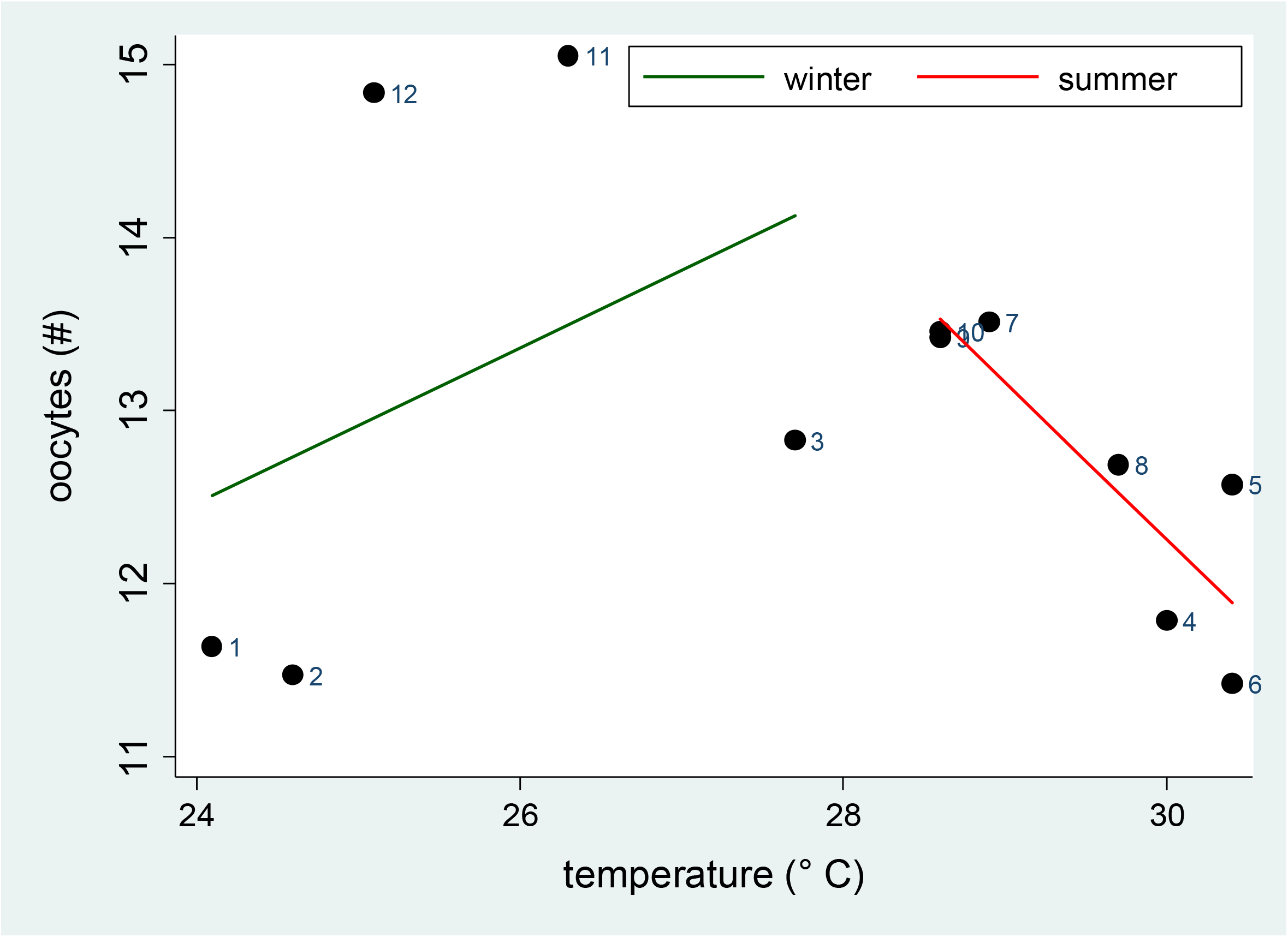
Monthly ambient temperature and # of oocytes retrieved.

Time-lagged exposure to ambient temperature, heat stress and temperature SD showed significant associations with the number of oocytes retrieved, especially when ambient temperatures were low and high, i.e., 1 SD below and 1 SD above the monthly average temperatures, respectively. For example, when the 1-week lag average temperature was below 25° C, 6 and 7-day lag weekly temperatures and heat stress showed statistically significant associations with poor quality oocytes (see Table S1 in the supplement online material (SOM)). When the average weekly temperature was greater than or equal to 30° C, the number of oocytes retrieved, and M1 and poor (or GV) grade oocytes showed significant associations with weekly lagged ambient temperature (Table S1 in SOM).

When adjusted for confounders, a week-lag temperature showed a significant inverse association with the number of oocytes retrieved, i.e. a unit increase in the week-lagged ambient temperature was associated with 1% decline in the number of oocytes. However, under hypothermic conditions (i.e. one-week lagged temperature was < 25° C), a unit increase in a week lag ambient temperature was associated with an 11% increase in the number of oocytes after adjusting for potential confounders (Table 4). SD of temperature under hypothermic condition showed an inverse association with the number of oocytes, i.e. one SD increase in a week-lag hourly temperature was associated with a 36% decrease in the number of oocytes retrieved (incidence rate ratio (IRR) = 0.64; 95% CI = 0.51 – 0.81; p < 0.01) (Table 5). Under hyperthermic conditions when a week-lagged temperature was >= 30° C, a unit increase in the weekly average temperature was associated with an 8% decrease in the number of oocytes (IRR = 0.92; 95% CI = 0.92 – 0.98; p < 0.01). Under hyperthermic conditions, temperature of SD showed a positive association with number of oocytes, i.e. with one 1SD increase (that refers to a sudden increase or decrease) in ambient temperature, number of oocytes retrieved increased by 81% per cent (IRR = 1.81; 95% CI = 1.30 to 2.52; p < 0.001).

**Table 4:** Association between # oocytes retrieval and a week lagged ambient temperature.

| Variables | Temperature thresholds |  |  |  |
| --- | --- | --- | --- | --- |
|  | All | < 25° C | 25° C – 30° C | >= 30° C |
| Age (years) | 0.98*** | 0.97*** | 0.97*** | 0.99 |
|  | (0.97 - 0.98) | (0.96 - 0.98) | (0.97 - 0.98) | (0.98 - 1.00) |
| AMH (ng/ml) | 1.07*** | 1.09*** | 1.07*** | 1.07*** |
|  | (1.06 - 1.08) | (1.07 - 1.11) | (1.06 - 1.08) | (1.05 - 1.08) |
| HMG dose (IU) | 1.00*** | 1 | 1.00*** | 1.00* |
|  | (1.00 - 1.00) | (1.00 - 1.00) | (1.00 - 1.00) | (1.00 - 1.00) |
| 1-7 d lag ambient temperature (° C) | 0.99** | 1.11** | 0.97*** | 0.92** |
|  | (0.98 - 1.00) | (1.02 - 1.21) | (0.95 - 0.99) | (0.87 - 0.98) |
| Constant | 25.64*** | 1.7 | 48.94*** | 148.07*** |
|  | (19.20 - 34.23) | (0.21 - 13.61) | (27.40 - 87.42) | (19.74 - 1,110.78) |
| Observations | 692 | 123 | 444 | 125 |
| *** p<0.01, ** p<0.05, * p<0.1 |  |  |  |  |

**Table 5:**
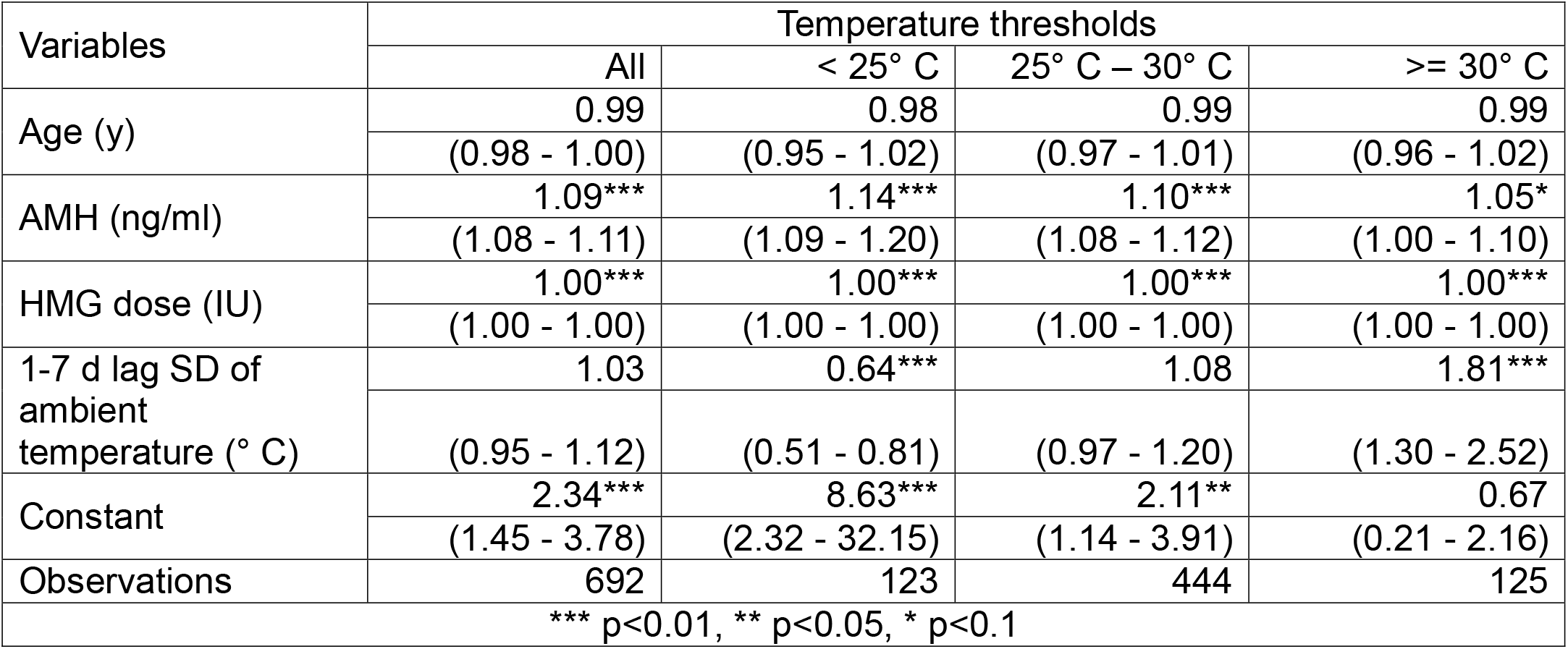
Association between # oocytes retrieval and a week lagged standard deviation of ambient temperature.

| Variables | Temperature thresholds |  |  |  |
| --- | --- | --- | --- | --- |
|  | All | < 25° C | 25° C – 30° C | >= 30° C |
| Age (y) | 0.99 | 0.98 | 0.99 | 0.99 |
|  | (0.98 - 1.00) | (0.95 - 1.02) | (0.97 - 1.01) | (0.96 - 1.02) |
| AMH (ng/ml) | 1.09*** | 1.14*** | 1.10*** | 1.05* |
|  | (1.08 - 1.11) | (1.09 - 1.20) | (1.08 - 1.12) | (1.00 - 1.10) |
| HMG dose (IU) | 1.00*** | 1.00*** | 1.00*** | 1.00*** |
|  | (1.00 - 1.00) | (1.00 - 1.00) | (1.00 - 1.00) | (1.00 - 1.00) |
| 1-7 d lag SD of ambient temperature (° C) | 1.03 | 0.64*** | 1.08 | 1.81*** |
|  | (0.95 - 1.12) | (0.51 - 0.81) | (0.97 - 1.20) | (1.30 - 2.52) |
| Constant | 2.34*** | 8.63*** | 2.11** | 0.67 |
|  | (1.45 - 3.78) | (2.32 - 32.15) | (1.14 - 3.91) | (0.21 - 2.16) |
| Observations | 692 | 123 | 444 | 125 |
| *** p<0.01, ** p<0.05, * p<0.1 |  |  |  |  |

When analyzing the impact of high temperatures, it is crucial to consider age and ovarian reserve, as indicated by the biomarker AMH. One week lagged ambient temperature showed a significant association with age, indicating that each additional year of age was associated with a 2% decrease in the number of oocytes retrieved (odds ratio = 0.98; 95% CI = 0.97 to 0.98; p < 0.01) across all temperature conditions. This effect was significant for temperatures below 25°C and between 25°C and 30°C. However, at temperatures above 30°C, age did not show a statistically significant association with the number of oocytes (Table 4).

For a week-lagged ambient temperature, each unit increase in AMH levels was associated with a 7% increase in the number of oocytes retrieved across all temperatures (odds ratio = 1.07; 95% CI = 1.06 – 1.08; p<0.01) and this effect was more pronounced below 25°C (Table 4). With a week-lagged standard deviation of ambient temperature, each unit increase in AMH levels was significantly associated with a 9% increase in the number of oocytes retrieved at all temperatures (odds ratio = 1.09; 95% CI = 1.08 and 1.11; p < 0.01), with a slight increase of 14% when the temperature was less than 25°C (Table 5). The HMG dose did not show a significant association with the number of oocytes retrieved across all temperature ranges for both the week-lagged ambient temperature and its standard deviation (Table 4 and Table 5).

#### Embryo and pregnancy confirmation

978 embryo transfer (ET) procedures were attempted across 690 patients. For 337 (34.64%) attempts, fresh embryos were transferred and for another 650 (65.54%) frozen embryos were used. Of the 978 ETs, 566 (57.87%) were incubated for three days, 377 (38.55%) for 5 days, and remaining 35 for 2 or 4 days. On average, ET was attempted 1.42 times per patient. Of the 978 ET attempts, 319 (32.62%) resulted in pregnancy confirmation 14 days after ET using beta HCG testing, and 249 (25.46%) resulted in clinical pregnancy confirmed four weeks after ET using transvaginal sonography. Average pregnancy confirmation rate was 27.0%, the highest rate (40.3%) occurring in May and the lowest (19.4%) in July.

We used two measures of the success of pregnancy: β-HCG testing two weeks after ET and ultrasound evaluation four weeks after ET. Given the reliability of ultrasound evaluation, the main results are restricted to ultrasound confirmation of pregnancy, and the results of β-HCG evaluation are presented in the Supplementary online Material (SOM) (Table – S5).

Overall, results show that embryo stage, number of good quality embryos and number of attempts showed significant associations with the clinical confirmation of pregnancy. Four or more days older embryos had 2.93 times higher odds of clinical confirmation of pregnancy as compared to 3-day old embryos (odds ratio = 2.93; 95 CI = 2.02 to 4.26; p < 0.001). Likewise, with a unit increase in good quality embryo, chance of clinical pregnancy confirmation increased by 51% (odds ratio = 1.51; 95% CI = 1.26 to 1.81; p < 0.001) (Table 6). Overall ambient temperature exposure did not show any significant association with pregnancy confirmation due to seasonality (Figure 2). Based on exploratory analysis, we found average ambient temperature and variations in temperature a week prior to ET and when temperature was ≥ 30° C three days before and after day of embryo transplantation showed the strongest inverse association with pregnancy confirmation.

**Table 6:** Association between clinical confirmation of pregnancy (1=yes, 0 otherwise) and selected covariates and exposure to a week-lagged ambient temperature and its variation.

| Variable name | Without ambient temperature exposure | 1-week lag Temperature (all observations) | Weekly Temperature $\geq 30^{\circ}\text{C} \pm 3\text{d}$ Embryo Transfer Day | | |
| --- | --- | --- | --- | --- | --- |
| | | | Mean 1 w-lag Temperature ($^{\circ}\text{C}$) | SD of 1 w-lag Temperature ($^{\circ}\text{C}$) | Mean X SD of 1w lag Temperature ($^{\circ}\text{C}$) |
| Embryo stage (1=3 d or more, 0 otherwise) | 2.93*** | 2.93*** | 2.14* | 2.60** | 2.66** |
|  | (2.02 – 4.26) | (2.01 – 4.26) | (0.90 – 5.13) | (1.06 – 6.41) | (1.07 – 6.57) |
| Embryo status (1 = frozen, 0 otherwise) | 1.18 | 1.21 | 5.23*** | 4.22** | 4.28** |
|  | (0.77 – 1.80) | (0.79 – 1.86) | (1.55 – 17.65) | (1.24 – 14.35) | (1.25 – 14.61) |
| Number of good quality embryos | 1.51*** | 1.50*** | 1.32 | 1.45 | 1.45 |
|  | (1.26 – 1.81) | (1.25 – 1.80) | (0.85 – 2.06) | (0.93 – 2.28) | (0.92 – 2.28) |
| Age (year) | 1.01 | 1.02 | 0.95 | 0.96 | 0.96 |
|  | (0.98 – 1.05) | (0.98 – 1.05) | (0.87 – 1.03) | (0.89 – 1.04) | (0.88 – 1.04) |
| Body Mass Index (weight (kg)/height ($\text{m}^2$)) | 1 | 1 | 1 | 1 | 1 |
|  | (1.00 – 1.00) | (1.00 – 1.00) | (0.97 – 1.03) | (0.96 – 1.03) | (0.97 – 1.03) |
| AMH dose (ng/ml) | 0.96 | 0.96 | 0.93 | 0.94 | 0.94 |
|  | (0.91 – 1.01) | (0.91 – 1.02) | (0.81 – 1.07) | (0.82 – 1.09) | (0.81 – 1.08) |
| Number of attempts of IVF treatment | 1.56*** | 1.55*** | 1.22 | 1.23 | 1.24 |
|  | (1.25 – 1.94) | (1.24 – 1.93) | (0.74 – 2.01) | (0.74 – 2.06) | (0.74 – 2.08) |
| 1-week lagged ambient temperature (one week prior to embryo transfer) |  | 0.98 | 0.61*** | 0.29*** | 0.96*** |
|  |  | (0.93 – 1.05) | (0.43 – 0.88) | (0.13 – 0.65) | (0.94 – 0.98) |
| Observations | 941 | 932 | 168 | 168 | 168 |

\*\*\* p<0.01, \*\* p<0.05, \* p<0.1
Note: clinical confirmation of pregnancy using ultrasound.

**Figure 2:**
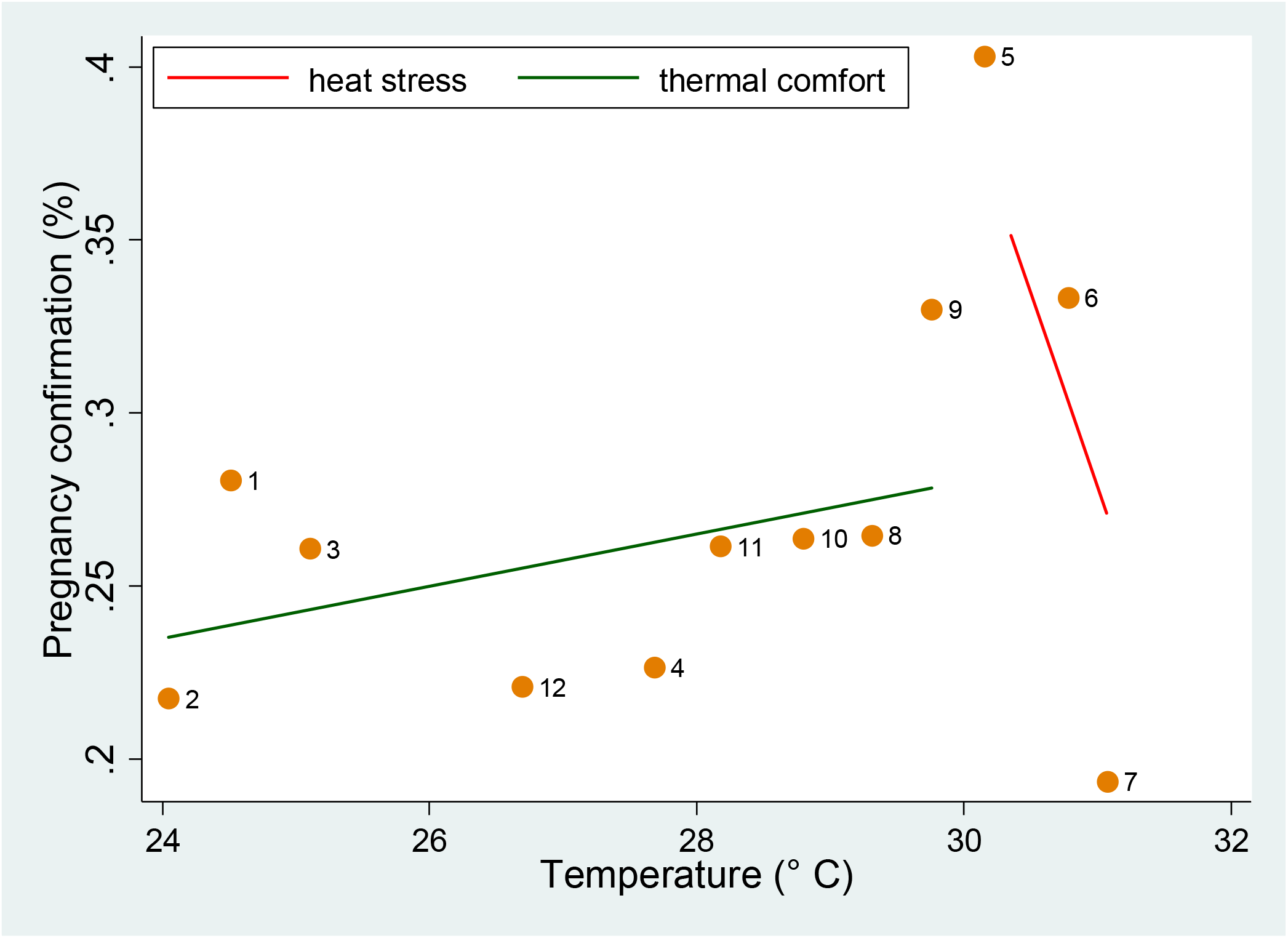
Monthly ambient temperature and pregnancy confirmation rate.

Under hyperthermic conditions, a unit increase in ambient temperature was associated with 39% lower odds of pregnancy confirmation (odds ratio = 0.61; 95% CI = 0.43 to 0.88; p < 0.01). Change in ambient temperature, i.e. sudden increase and/or decrease significantly reduced the odds of pregnancy confirmation, e.g. with a SD increase in a week lagged temperature odds of pregnancy confirmation decreased by 71% (odds ratio = 0.29; 95% CI = 0.13 and 0.65; p < 0.01). Interaction of ambient temperature and variation in temperature significantly lowers the odds of pregnancy confirmation (Table 6).

#### Pregnancy outcomes

Of the confirmed clinical pregnancies, 117 (47%) resulted in full-term delivery, 19 (7.63%) in preterm birth, and the rest 136 (45.37%) in SAM.

#### Spontaneous abortion/Miscarriage (SAM)

Average SAM rate was 28.0% ranging from 12.5% in October to 45.5% in March (Table 3; Figure 3). We found a positive association between ambient temperature and SAM (Figure 4). After adjusting for confounders, a 10° F increase in two-day lag heat stress was associated with a 3% higher odds of SAM (odds ratio ~ 1.03; 95% CI = 1.001 to 1.068; p-value < 0.043); during high heat-stress months (April through October) there was 2.12 times higher odds of SAM as compared to winter months (odd ratio ~ 2.12; 95% CI = 1.14 - 3.94, p < 0.01) (Table 7).

**Table 7:** Spontaneous abortion with respect to time-lagged meteorological conditions (odds ratio; 95% confidence interval in parenthesis).

|  |  |  |  |  |
| --- | --- | --- | --- | --- |
| Heat index 2d lag | 1.03* |  | 1.03** |  |
|  | (1.00 - 1.06) |  | (1.00 - 1.07) |  |
| Season category (Mar-Aug, 1, 0) |  | 2.12** |  | 2.17** |
|  |  | (1.14 - 3.94) |  | (1.13 - 4.19) |
| Age (y) |  |  | 1.02 | 1.01 |
|  |  |  | (0.94 - 1.10) | (0.94 - 1.10) |
| BMI (weight kg/height(m<sup>2</sup>)) |  |  | 1.04 | 1.03 |
|  |  |  | (0.95 - 1.13) | (0.94 - 1.12) |
| HMG (IU) |  |  | 1.00*** | 1.00*** |
|  |  |  | (1.00 - 1.00) | (1.00 - 1.00) |
| AMH (ng/ml) |  |  | 1.06 | 1.05 |
|  |  |  | (0.95 - 1.19) | (0.93 - 1.17) |
| Number of IVF tries |  |  | 1.4 | 1.44* |
|  |  |  | (0.92 - 2.14) | (0.94 - 2.21) |
| Constant | 0.02** | 0.26*** | 0.00*** | 0.01*** |
|  | (0.00 - 0.42) | (0.16 - 0.41) | (0.00 - 0.07) | (0.00 - 0.35) |
| Observations | 211 | 211 | 211 | 211 |
| *** p<0.01, ** p<0.05, * p<0.1 |  |  |  |  |

**Figure 3:**
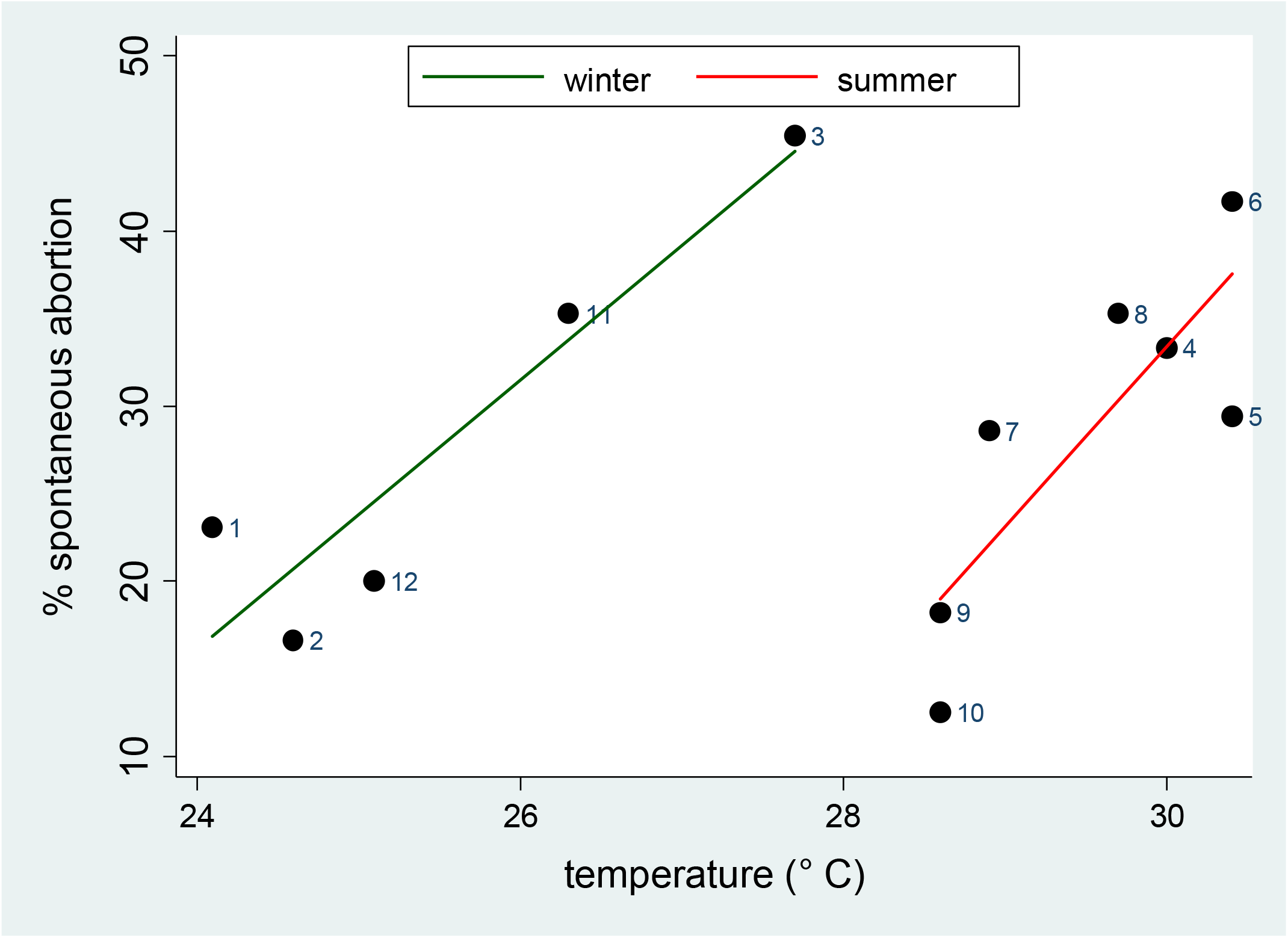
Monthly ambient temperature and spontaneous abortion rate.

#### PTB and heat stress

Of all clinical pregnancies, 19 resulted in PTB and 14 of them occurred in months when monthly average temperature was > 28° C and heat index > 90° F (or 32.2°C) (Table 8). We modelled the risk of PTB with respect to ambient temperature, heat index and their SD. Although the frequency of PTB was relatively higher when time-lagged temperatures, HI and their SD were higher, they were statistically insignificant. However, the risks of PTB increased significantly under extreme hyperthermic conditions. For example, odds of PTB were three times higher when three-day lagged HI was greater than 95° F (or > 35° C) as compared to when 3d lag HI was < 95° F (Table 7). When adjusted for confounders, PTB odds were 3.97 times higher under hyperthermic conditions (odds ratio = 3.97, 95% CI = 1.35 to 11.65, p < 0.05). Number of IVF tries was also associated with lower odds of PTB, e.g. with every IVF try PTB odds declined more than 50% (odds ratio = 0.41; 95% CI = 0.19 to 0.91; p < 0.05).

**Table 8:** PTB with respect to time-lagged meteorological conditions (odds ratio; 95% confidence interval in parenthesis)

|  | 3d lag<br>HI | HI Cat<br>(>95<br>=1,0) | Season<br>(1=3 to 10,<br>0<br>otherwise) | 3d lag HI | 3d lag HI ><br>95 | Season<br>(1=3 to 10, 0<br>otherwise) |
| --- | --- | --- | --- | --- | --- | --- |
| Exposure<br>measure | 1.042<br>(0.988 -<br>1.100) | 3.006**<br>(1.131 -<br>7.993) | 1.413<br>(0.448 -<br>4.454) | 1.048<br>(0.990 -<br>1.109) | 3.970**<br>(1.353 -<br>11.647) | 1.56<br>(0.459 -<br>5.297) |
| Age (years) |  |  | 1<br>(0.91 -<br>1.09) | 0.996<br>(0.910 -<br>1.091) | 0.997<br>(0.906 -<br>1.097) | 0.989<br>(0.905 -<br>1.081) |
| BMI (weight<br>kg/height(m<sup>2</sup>)) |  |  | 0.97<br>(0.87 -<br>1.09) | 0.975<br>(0.868 -<br>1.095) | 0.988<br>(0.878 -<br>1.111) | 0.983<br>(0.882 -<br>1.096) |
| HMG (IU) |  |  | 0.998***<br>(1.00 -<br>1.00) | 0.999***<br>(0.998 -<br>0.999) | 0.999***<br>(0.998 -<br>0.999) | 0.999***<br>(0.998 -<br>0.999) |
| AMH (ng/ml) |  |  | 0.91<br>(0.75 -<br>1.11) | 0.912<br>(0.750 -<br>1.108) | 0.89<br>(0.724 -<br>1.094) | 0.918<br>(0.758 -<br>1.111) |
| Number of IVF<br>tries |  |  | 0.46**<br>(0.21 -<br>0.99) | 0.460**<br>(0.215 -<br>0.986) | 0.413**<br>(0.188 -<br>0.910) | 0.473**<br>(0.224 -<br>0.999) |
| Constant | 0.002**<br>(0.000 -<br>0.336) | 0.058***<br>(0.027 -<br>0.124) | 0.077***<br>(0.028 -<br>0.213) | 0.06<br>(0.000 -<br>37.907) | 2.271<br>(0.029 -<br>175.989) | 2.917<br>(0.050 -<br>170.717) |
| Observations | 209 | 209 | 209 | 209 | 209 | 209 |
| *** p<0.01, ** p<0.05, * p<0.1 |  |  |  |  |  |  |

HI: Heat Index

## D. DISCUSSION

Using comprehensively abstracted clinical chart data with time stamp for each IVF procedure, our multi-site study found that exposure to both thermal discomfort (very low and high temperatures) and thermal shock due to sudden changes in temperature were significantly associated with IVF outcomes. Specifically, under both hypothermic and hyperthermic conditions and sudden changes in ambient temperature significantly reduced the quantity and quality of oocytes. Under hyperthermic conditions (i.e. > 30° C) increase in ambient temperature and sudden changes in ambient temperature a few days before and after ET significantly reduced the pregnancy rates. This finding sheds a novel insight into how pre- and post-ET heat-stress and thermal shock diminish the likelihood of IVF assisted pregnancy. Among these two measures, thermal shock emerged as a more important predictor of pregnancy rate than the exposure to ambient temperature. Our analysis also showed that both heat stress and seasonality were significant predictors of SAM and PTB.

Some of our findings are consistent with the previous epidemiological studies that show significant associations between IVF outcomes and exposure to ambient temperature and seasonality.^35^ Although toxicological studies document mechanisms underlying the effects of thermal discomfort on IVF outcomes, several studies did not find any significant associations between thermal discomfort and IVF outcomes.^36^ For example, studies conducted in Israel and China did not show any significant associations between ambient temperature and oocytes quantity.^26,27^ Likewise, a nationwide study in US found no association between IVF cycles and IVF outcomes except in the Western parts of the US.^37^ As detailed in section “A” there are several reasons for inconsistencies in the literature, including lack of location- and time-stamped exposure data, variations in the physiological and pathological conditions of both patients and their partners, control for confounders and region-specific variations in adaptation to thermal discomfort.

*In-vitro* and *in-vivo* studies substantiate the causal mechanisms underlying the effects of thermal discomfort and thermal shock on IVF outcomes. Core body temperature is sensitive to exposure to internal and external heat- and cold-stress.^38,39^ When the core temperature exceeds or drops below hypothalamic set point peripheral and thermal receptors are activated which signal thermoregulatory processes to compensate for either loss or gain of core body temperature.^40^ These processes directly affect cardiac output, metabolic rate, nutrient delivery, electrolyte balance and hormonal disruption. For example, under hyperthermic conditions, cooling mechanism is activated, increasing the sweat secretion by increasing peripheral blood flow to dissipate heat through radiation and evaporative cooling.^41^ This compromises blood flow, nutrient delivery and electrolyte imbalance in critical organs, including heart, kidney and fetus. Thus prolonged heat exposure can induce heat shock proteins, oxidative stress and initiate a cascade of inflammatory processes, including cellular changes and organ damage.^42^ Cold stress also affect reproductive health by inducting inflammatory cytokines and differential gene expression that affect hormonal changes as well as oxidative stress pathways in both uterus and ovaries.^43–45^ Sudden changes in temperature that are often accompanied by changes in barometric pressure are widely recognized for causing weather-related illnesses by thermal shock and stressing immune response,^46^ because of body’s narrow thermoelasticity to compensate for excess loss/gain heat due to abrupt decline and rise in temperature. Literature suggest that both thermal discomfort and thermal shock affect both oogenesis and pregnancy, especially in early stages of pregnancy.^47^ For example, early-stage embryos, i.e. 1-to 8-cell embryos, are highly susceptible to heat- and cold-stress,^47^ as embryonic gene and protein activation, including HSP, occurs ~ 3d after fertilization.^48^

Our findings concerning the association between adverse IVF outcomes and cold- and heat-stress and thermal shock are supported by the established biological mechanisms. These findings have important clinical-translational and public health implications. First, patients’ thermal discomfort exposure days and weeks prior to each IVF procedure must be carefully evaluated and healthcare must potentially screen patients for heat shock proteins, oxidative stress and inflammation. Second, patients must be protected from exposure to thermal discomfort and thermal shock for at least a week prior to embryo transfer and throughout the pregnancy. Third, the findings of this research must be integrated in clinical training so that healthcare providers can engage their patients in accessing publicly accessible weather data and strategies to mitigate and/or avoid their exposure to thermal discomfort and thermal shock.

Although our paper provides novel insight into the role of thermal discomfort and thermal shock in IVF outcomes, our findings had some limitations. First, patients in the study have various underlying causes of infertility and coexisting medical conditions, which may potentially influence the results of IVF treatment.Second, even though ICSI was standardized and performed during all IVF cycles to control for male factors including severe oligoasthenoteratozoospermia (OATS), we cannot completely rule out the effects of thermal distress related DNA fragmentation that can affect both quality and quantity of the embryos and thus pregnancy outcomes. Third, we did not consider the impact of other environmental stressors, such as exposure to environmental pollutants, especially endocrine disruptors through inhalation, ingestion and dermal routes which can also impact IVF outcomes. Fourth, this study relied on ambient heat- and cold-stress. However, exposure to heat- and cold-stress and/or thermal comfort in home- and work environments can confound each stage of IVF treatment and outcomes. Fifth, although our study includes three IVF centers located in the southern parts of India, which are prone to chronic heat stress. This limits the generalizability of our findings to other parts of the world, including temperate and polar regions, where intensity, frequency and duration of both heat- and cold-stress and thermal shock can be very different. Sixth, some of the IVF procedures were conducted during the COVID-19 pandemic, which might have limited the frequency of IVF procedures and number of patients during the pandemic. Finally, some patients dropped out of our IVF clinics after pregnancy confirmation, and our study lacked access to all pregnancy outcomes. Given the above limitations, our results on the role of thermal discomfort, thermal shock and seasonality in IVF outcomes must be interpreted in the light of these limitations.

## Conclusions and recommendations

Given declining fertility rate and increasing demand for IVF, our research systematically documented the role of thermal discomfort, thermal shock and seasonality in IVF outcomes ranging from quantity and quality of oocytes to pregnancy outcomes. These findings, especially in areas and seasons, which are prone to heat- and cold-stress and sudden changes in temperature, warrant attention to protect patients from exposure to thermal discomfort and thermal shock before and during all stages of IVF. These findings warrant translation into clinical practice so that fertility clinicians can train and engage their patients’ exposure mitigation and avoidance strategies to improve the likelihood of IVF outcomes. Some of these strategies could be as simple as avoiding working during peak temperatures of the day and accessing climate-controlled environments at the workplace for individuals undergoing fertility treatments. Additionally, fertility specialists may explore seasonal timing of treatment cycles and adjunct interventions such as prescribing mitochondrial nutrients like L-carnitine, CoQ10, and antioxidants to reduce systemic inflammation and ROS to optimize outcomes in warmer climates.

## Data Availability

All de-identified data of the study can be made available upon a reasonable request for analysis and/or re-analysis.

