## Supplement Tables for "Thermal Discomfort and *In-vitro* Fertilization Treatment Outcomes"

**Supplementary online material**

**Table S1 – Pairwise correlation between meteorological conditions and oocyte retrieval when 1-7 d lagged temperature**

**was below 25° C (i.e. 1 SD below the annual average temperature).**

|  |  | # of oocytes retrieved | % very good quality oocytes | % good quality oocytes | % poor quality oocytes |
| --- | --- | --- | --- | --- | --- |
| Number of oocytes retrieved | Total | 1 |  |  |  |
|  | M1 grade | 0.7624* | 1 |  |  |
|  | M2 grade | 0.2653* | -0.3371* | 1 |  |
|  | GV grade | 0.4870* | 0.2916* | -0.0758* | 1 |
| Time-lagged average ambient temperature (° C) | 1 d | -0.0659 | -0.0428 | -0.0559 | -0.0349 |
|  | 2 d | -0.0456 | -0.0371 | -0.0309 | -0.0384 |
|  | 3 d | -0.0589 | -0.0281 | -0.0489 | -0.0681 |
|  | 4 d | -0.0622 | -0.0363 | -0.0542 | -0.0287 |
|  | 5 d | -0.0289 | 0.0001 | -0.0561 | -0.0089 |
|  | 6 d | -0.0001 | 0.036 | -0.0748* | 0.0214 |
|  | 7 d | 0.0039 | 0.0322 | -0.0682 | 0.0444 |
|  | 1-7 day | -0.0393 | -0.0116 | -0.0594 | -0.0172 |
| Time-lagged ambient heat stress exposure (° F) | 1 d | -0.0611 | -0.0259 | -0.0701 | -0.0295 |
|  | 2 d | -0.0447 | -0.0245 | -0.0478 | -0.0246 |
|  | 3 d | -0.0626 | -0.0197 | -0.065 | -0.0588 |
|  | 4 d | -0.064 | -0.0316 | -0.0617 | -0.0237 |
|  | 5 d | -0.0312 | 0.0041 | -0.0658 | -0.0043 |
|  | 6 d | -0.0122 | 0.0279 | -0.0738 | 0.0034 |
|  | 7 d | -0.0068 | 0.0259 | -0.0698 | 0.0302 |
|  | 1-7 day | -0.0428 | -0.0066 | -0.0689 | -0.0162 |
| Time-lagged standard deviation of hourly ambient temperature (° C) | 1 d | -0.0089 | -0.0604 | 0.0426 | 0.0217 |
|  | 2 d | 0.044 | -0.0019 | 0.0224 | 0.03 |
|  | 3 d | 0.0244 | 0.0002 | 0.0203 | -0.0285 |
|  | 4 d | -0.0006 | -0.0205 | 0.0096 | -0.0014 |
|  | 5 d | -0.0131 | -0.0265 | 0.0006 | 0.0203 |
|  | 6 d | 0.0001 | 0.0105 | -0.0366 | 0.0283 |
|  | 7 d | -0.0175 | -0.0212 | -0.0216 | 0.0503 |
|  | 1-7 day | 0.0048 | -0.0207 | 0.0061 | 0.0211 |

* p < 0.05

**Table S2 – Pairwise correlation between meteorological conditions and oocyte retrieval when 1-7 d lagged temperature was below 25° C and 30° C (i.e. 1 SD below the annual average temperature).**

|  |  | # of oocytes retrieved | % very good quality oocytes | % good quality oocytes | % poor quality oocytes |
| --- | --- | --- | --- | --- | --- |
| Number of oocytes retrieved | Total | 1 |  |  |  |
|  | M1 grade | 0.7571* | 1 |  |  |
|  | M2 grade | 0.2755* | -0.3490* | 1 |  |
|  | GV grade | 0.4774* | 0.3019* | -0.0787 | 1 |
| Time-lagged average ambient temperature (° C) | 1 d | -0.0631 | -0.0817 | -0.0093 | -0.0132 |
|  | 2 d | -0.0344 | -0.0805 | 0.0294 | -0.0294 |
|  | 3 d | -0.0716 | -0.0681 | -0.016 | -0.0962* |
|  | 4 d | -0.0725 | -0.0974* | 0.006 | -0.0262 |
|  | 5 d | -0.0063 | -0.0224 | -0.0023 | 0.0126 |
|  | 6 d | 0.044 | 0.0381 | -0.032 | 0.0502 |
|  | 7 d | 0.0461 | 0.0272 | -0.0163 | 0.063 |
|  | 1-7 day | -0.028 | -0.05 | -0.0066 | -0.0073 |
| Time-lagged ambient heat stress exposure (° F) | 1 d | -0.057 | -0.0597 | -0.0298 | -0.0002 |
|  | 2 d | -0.0343 | -0.063 | 0.01 | -0.0091 |
|  | 3 d | -0.0544 | -0.0406 | -0.0311 | -0.0695 |
|  | 4 d | -0.0572 | -0.0727 | -0.0037 | -0.0211 |
|  | 5 d | -0.0057 | -0.0184 | -0.0141 | 0.0211 |
|  | 6 d | 0.026 | 0.0269 | -0.0331 | 0.0266 |
|  | 7 d | 0.0363 | 0.0282 | -0.0253 | 0.0462 |
|  | 1-7 day | -0.0235 | -0.0321 | -0.0208 | -0.0004 |
| Time-lagged standard deviation of hourly ambient temperature (° C) | 1 d | -0.0242 | -0.0597 | 0.038 | -0.0144 |
|  | 2 d | 0.0438 | 0.0015 | 0.0257 | 0.0279 |
|  | 3 d | 0.0124 | 0.0005 | 0.0123 | -0.0491 |
|  | 4 d | -0.0223 | -0.0416 | 0.0167 | -0.0107 |
|  | 5 d | -0.0091 | -0.0274 | 0.0243 | 0.0197 |
|  | 6 d | 0.0117 | 0.006 | -0.0114 | 0.0467 |
|  | 7 d | -0.0235 | -0.0229 | -0.0258 | 0.0594 |
|  | 1-7 day | -0.0021 | -0.0245 | 0.0133 | 0.014 |

* p < 0.05

**Table S4 – Pairwise correlation between meteorological conditions and oocyte retrieval when**

**1-7 d lagged temperature was above 30° C (i.e. 1 SD below the annual average temperature).**

|  |  | # of oocytes retrieved | % very good quality oocytes | % good quality oocytes | % poor quality oocytes |
| --- | --- | --- | --- | --- | --- |
| Number of oocytes retrieved | Total | 1 |  |  |  |
|  | M1 grade | 0.8064* | 1 |  |  |
|  | M2 grade | 0.1423 | -0.3459* | 1 |  |
|  | GV grade | 0.4230* | 0.2299* | -0.1177 | 1 |
| Time-lagged average ambient temperature (° C) | 1 d | -0.2628* | -0.3112* | 0.0919 | -0.168 |
|  | 2 d | -0.2091* | -0.2452* | 0.054 | -0.0785 |
|  | 3 d | -0.1935* | -0.2369* | 0.1213 | -0.1805* |
|  | 4 d | -0.1511 | -0.2072* | 0.0819 | -0.0113 |
|  | 5 d | -0.0737 | -0.1191 | 0.0379 | 0.1012 |
|  | 6 d | -0.079 | -0.0642 | 0.0008 | 0.0529 |
|  | 7 d | -0.0209 | -0.0407 | 0.0114 | 0.2234* |
|  | 1-7 day | -0.2028* | -0.2515* | 0.0822 | -0.0032 |
| Time-lagged ambient heat stress exposure (° F) | 1 d | -0.1865* | -0.1820* | 0.0351 | -0.1781* |
|  | 2 d | -0.1441 | -0.1389 | -0.0212 | -0.051 |
|  | 3 d | -0.2400* | -0.2229* | 0.052 | -0.1513 |
|  | 4 d | -0.2176* | -0.2126* | -0.001 | 0.0198 |
|  | 5 d | -0.0906 | -0.0671 | -0.0358 | 0.0782 |
|  | 6 d | -0.1034 | -0.0851 | 0.0107 | 0.0277 |
|  | 7 d | -0.0918 | -0.0947 | -0.0066 | 0.1777* |
|  | 1-7 day | -0.2140* | -0.2005* | 0.0051 | -0.0086 |
| Time-lagged standard deviation of hourly ambient temperature (° C) | 1 d | -0.0771 | -0.1888* | 0.0855 | 0.0852 |
|  | 2 d | -0.0532 | -0.1503 | 0.0413 | 0.0874 |
|  | 3 d | -0.0129 | -0.0759 | 0.0239 | 0.068 |
|  | 4 d | 0.0278 | -0.0623 | 0.0418 | 0.1581 |
|  | 5 d | -0.0317 | -0.0914 | -0.0469 | 0.1644 |
|  | 6 d | -0.1133 | -0.1145 | -0.0814 | 0.1004 |
|  | 7 d | -0.0374 | -0.0833 | -0.0259 | 0.1885* |
|  | 1-7 day | -0.0605 | -0.1587 | 0.0062 | 0.1823* |

* p < 0.05

**Table S4: Number of oocytes with respect to 31 to 37 day lag SD of daily hourly temperature**

**(incidence risk ratio; 95% confidence interval in parentheses).**

| Thermal comfort (25°C-29°C = 1, 0 otherwise | 1.07*** |  |  |  | 1.08*** |
| --- | --- | --- | --- | --- | --- |
|  | (1.03 - 1.10) |  |  |  | (1.05 - 1.12) |
| SD temperature 31-37d lag |  | 0.92*** |  |  | 0.94*** |
|  |  | (0.90 - 0.94) |  |  | (0.92 - 0.97) |
| Age (years) |  |  |  |  | 0.98*** |
|  |  |  |  |  | (0.97 - 0.98) |
| AMH (ng/ml) |  |  |  |  | 1.03*** |
|  |  |  |  |  | (1.03 - 1.03) |
| HMG dose (IU) |  |  |  |  | 1.00*** |
|  |  |  |  |  | (1.00 - 1.00) |
| Temperature (° C) |  |  | 0.98*** |  |  |
|  |  |  | (0.98 - 0.99) |  |  |
| Heat index 4d lag (°F) |  |  |  | 1.00*** |  |
|  |  |  |  | (0.99 - 1.00) |  |
| Constant | 12.64*** | 15.97*** | 20.69*** | 17.84*** | 28.47*** |
|  | (12.36 - 12.93) | (15.09 - 16.90) | (17.38 - 24.63) | (15.24 - 20.89) | (25.14 - 32.24) |
| Observations | 1,106 | 1,104 | 1,104 | 1,104 | 1,104 |
| *** p<0.01, ** p<0.05, * p<0.1 | | | | | |

**Table S5. Association between biochemical pregnancy (1=yes, 0 otherwise) by testing serum β-HCG testing and selected covariates and exposure to a week-lagged ambient temperature and its variation.**

| Variable name | Without Temperature | 1-week lag Temperature (all observations) | Weekly Temperature ≥ 30° C ± 3d Embryo Transfer Day | | |
| --- | --- | --- | --- | --- | --- |
|  |  |  | Mean 1 w-lag Temperature (° C) | SD of 1 w-lag Temperature (° C) | Mean X SD of 1w lag Temperature (° C) |
| Embryo stage (1=3 d or more, 0 otherwise) | 2.29*** | 2.26*** | 2.53** | 3.01** | 3.08** |
|  | (1.62 - 3.24) | (1.59 - 3.20) | (1.06 - 6.00) | (1.23 - 7.36) | (1.25 - 7.57) |
| Embryo status (1 = frozen, 0 otherwise) | 0.98 | 1 | 3.72** | 3.10** | 3.13** |
|  | (0.66 - 1.45) | (0.67 - 1.48) | (1.28 - 10.84) | (1.04 - 9.20) | (1.05 - 9.30) |
| Number of good quality embryos | 1.62*** | 1.62*** | 1.60** | 1.75** | 1.75** |
|  | (1.37 - 1.92) | (1.37 - 1.91) | (1.04 - 2.46) | (1.13 - 2.71) | (1.13 - 2.71) |
| Age (years) | 1.02 | 1.02 | 0.98 | 1 | 0.99 |
|  | (0.99 - 1.05) | (0.99 - 1.05) | (0.91 - 1.06) | (0.92 - 1.08) | (0.92 - 1.08) |
| Body Mass Index (weight (kg)/height (m^2^)) | 1.02 | 1.02 | 1.05 | 1.05 | 1.05 |
|  | (0.99 - 1.06) | (0.99 - 1.06) | (0.98 - 1.13) | (0.97 - 1.13) | (0.97 - 1.13) |
| AMH ~~dose~~ (ng/ml) | 0.98 | 0.99 | 0.93 | 0.94 | 0.94 |
|  | (0.94 - 1.03) | (0.95 - 1.03) | (0.81 - 1.06) | (0.82 - 1.08) | (0.81 - 1.08) |
| Number of attempts of IVF treatment | 1.43*** | 1.42*** | 1.18 | 1.2 | 1.2 |
|  | (1.16 - 1.76) | (1.15 - 1.75) | (0.72 - 1.92) | (0.72 - 1.98) | (0.73 - 1.99) |
| 1-week lagged ambient temperature (one week prior to embryo transfer) |  | 0.99 | 0.63*** | 0.34*** | 0.97*** |
|  |  | (0.94 - 1.05) | (0.44 - 0.88) | (0.16 - 0.72) | (0.94 - 0.99) |
| Observations | 970 | 978 | 168 | 168 | 168 |

Note: Biochemical pregnancy using serum β-HCG**.**
